# The Diagnostic Potential of Combined Photoplethysmography and Ankle-Brachial Index in Peripheral Arterial Obstructive Disease: A Comparative Analysis with Doppler Ultrasound

**DOI:** 10.1101/2024.02.12.24302737

**Authors:** Han-Luen Huang, Chia-Hsiu Chang, Men-Tzung Lo, Chen Lin

## Abstract

**Background and aims:** While ankle-brachial index (ABI) and photoplethysmography (PPG) have also shown adequate sensitivity in detecting PAOD, their accuracies can be compromised in detecting milder cases. We aim to evaluate the diagnostic value of combining ABI and PPG in detecting PAOD for subjects with high atherosclerotic cardiovascular risks compared to duplex sonography.

**Methods:** 130 participants underwent ABI, PPG, and duplex sonography during the evaluation process. Two parameters were derived from the PPG-PPG amplitude ratio of the lower-to-upper extremities (PPG_ratio_) and the PPG amplitude of the lower extremity (PPG_amp_). Sensitivity, specificity, accuracy, and the area under receiver operative curve (AUC) were calculated for PPG parameters and ABI, and the combination of both methods. Multivariate analysis was performed for adjusting other relevant risk factors.

**Results:** 65 participants were diagnosed with PAOD based on duplex sonography. The ROC analysis revealed optimal cut-off values in diagnosing PAOD were 0.417 for PPG_ratio_ and 58 for PPG_amp_. Both PPG_ratio_ and PPG_amp_ demonstrated significantly higher sensitivities, 78.4% and 75.7%, in comparison to 55.9% of ABI<0.9 (p<0.05). Lower PPG_ratio_ and PPG_amp_ remained independently associated with PAOD even after adjusting for confounding factors. The AUC of combination models, including model 1 (ABI and PPG_ratio_), model 2 (ABI and PPG_amp_), and model 3 (ABI, PPG_ratio_, and PPG_amp_), exhibited improved performance with AUCs of 0.922, 0.922, and 0.931 (all p<0.01) compared to that of ABI alone (AUC:0.822).

**Conclusions:** The combination of ABI and PPG increases the sensitivity and accuracy prominently in diagnosing PAOD compared to ABI or PPG alone.

## Introduction

Peripheral Arterial Obstructive Disease (PAOD) stands as a significant cause of both morbidity and mortality within the spectrum of atherosclerotic cardiovascular diseases. This condition, which encompasses both symptomatic and asymptomatic patients, has garnered considerable attention due to its widespread prevalence. Epidemiological studies have consistently highlighted key risk factors associated with PAOD, including age, hypertension, diabetes mellitus, smoking, and dyslipidemia. [1, 2] While PAOD can manifest in symptoms such as intermittent claudication to even tissue loss, it often remains underdiagnosed, primarily due to the presence of asymptomatic cases and the unreliability of physical examination findings. [3, 4]

Early identification of peripheral arterial obstructive disease (PAOD) is crucial for prompt treatment and effective risk factor management. For early detection of these patients at high risk of atherosclerosis, the measurement of Ankle-Brachial Index (ABI) has been widely recommended as a first-line screening tool. [5-8] A lower ABI, indicating a decreased pressure ratio between ankle and brachial arteries, correlates with higher cardiovascular mortality. [9, 10] However, in a meta-analysis, Xu, D., et al. reported in patients with diabetes or advanced age, ABI has a high specificity but lower sensitivity (at best 80%) than that reported in earlier studies due to arterial calcification. [11] In such cases, further assessment using Toe-Brachial Index (TBI) in asymptomatic patients when ABI> 1.3 or treadmill exercise testing in symptomatic patients with normal ABI is advised. [12]

Photoplethysmography (PPG) emerges as a promising technique for assessing arterial pulsations in the lower extremities, and detecting perfusion abnormalities due to arterial stenosis or obstruction such as reduced pulse amplitude and delayed or dampened waveforms. Previous studies have demonstrated that PPG exhibits high sensitivity and specificity in detecting PAOD when compared to traditional ABI measurements. [13, 14] ABI and PPG employ different mechanisms for PAOD detection, yet they share similarities in measurement preparation. We hypothesize that PPG’s pulse amplitude direct evaluation of distal perfusion may compensate for ABI’s lower diagnostic sensitivity. To date, no study has systematically compared ABI, parameters derived from PPG, and combinations in diagnosing PAOD in subjects with high cardiovascular risk, utilizing duplex sonography as the reference standard.[15] This study aims to comprehensively assess the diagnostic value of ABI and PPG-derived parameters in individuals with high cardiovascular risk, based on results obtained from duplex sonography.

## Materials and methods

### Participants

This prospective observational study was conducted at the Cardiovascular Center of Hsinchu Cathay General Hospital. Between October 24, 2019, and October 12, 2021, we prospectively enrolled a total of 130 participants, from both the outpatient department and ward, with high atherosclerotic cardiovascular risks or showed suspicion of PAOD and had been recommended for ABI evaluation. Patients with uremia or those who cannot receive photoplethysmography examination were excluded. The examination sequence involved an initial ABI assessment, followed by PPG examination and duplex sonography. Demographic data, including age, sex, height, weight, Body Mass Index (BMI), and a comprehensive review of cardiovascular risk factors (such as diabetes mellitus with varying durations, hypertension, dyslipidemia, and smoking habits), as well as cardiovascular comorbidities, were extracted from electronic medical charts. All the collected data was approved by the institutional review board of the Cathay General Hospital (CGH-P108042).

### Experimental Design and Setup

Participants were instructed to lie flat on the examination table in a room maintained at a temperature of at least 23°C for a 5-minute resting period before undergoing both the Ankle-Brachial Index (ABI) and photoplethysmography (PPG) assessments. A single technician conducted both tests using a peripheral vascular diagnosis system (FALCON/PRO, VIASONIX), equipped with an 8 MHz continuous-wave Doppler probe and a PPG system. Subsequently, participants underwent duplex sonography using an ultrasound system and a 3-11 MHz continuous-wave Doppler probe (IE33, Phillips, USA), performed by a single cardiologist on the same examination table at room temperature. An infrared heater had been temporarily used to keep the room temperature above 23 °C if the initial room temperature fell below this threshold. The average temperature was 24.4±0.7 °C, with an average humidity was 55.1±4.1 %rh at the end of the PPG measurement.

### Ankle-Brachial Pressure Index Measurement and Photoplethysmography-derived Parameters

Automatic measurements of ABI were taken for both brachial arteries, posterior tibial arteries (PTA), and dorsalis pedis arteries (DPA). The numerator represented the higher systolic pressure between PTA and DPA for each lower extremity, while the denominator was determined by the higher of the two brachial artery systolic pressures. An abnormal ABI was defined as less than 0.9.

The PPG system employed a transmittance-type PPG clip to measure light absorption through amplitude differences between an emitter and a photodetector, sampled at 32 Hz. Each clip featured two light-emitting diodes (LEDs) emitting light at wavelengths of 700 nm and 840 nm. Four PPG clips were attached to the distal extremities of the participants, specifically on both middle fingers and second toes (Figure 1). PPG waveforms from all four extremities were simultaneously recorded for a minimum of 5 minutes. Two primary parameters were derived from the PPG waveforms:

**Figure 1.**
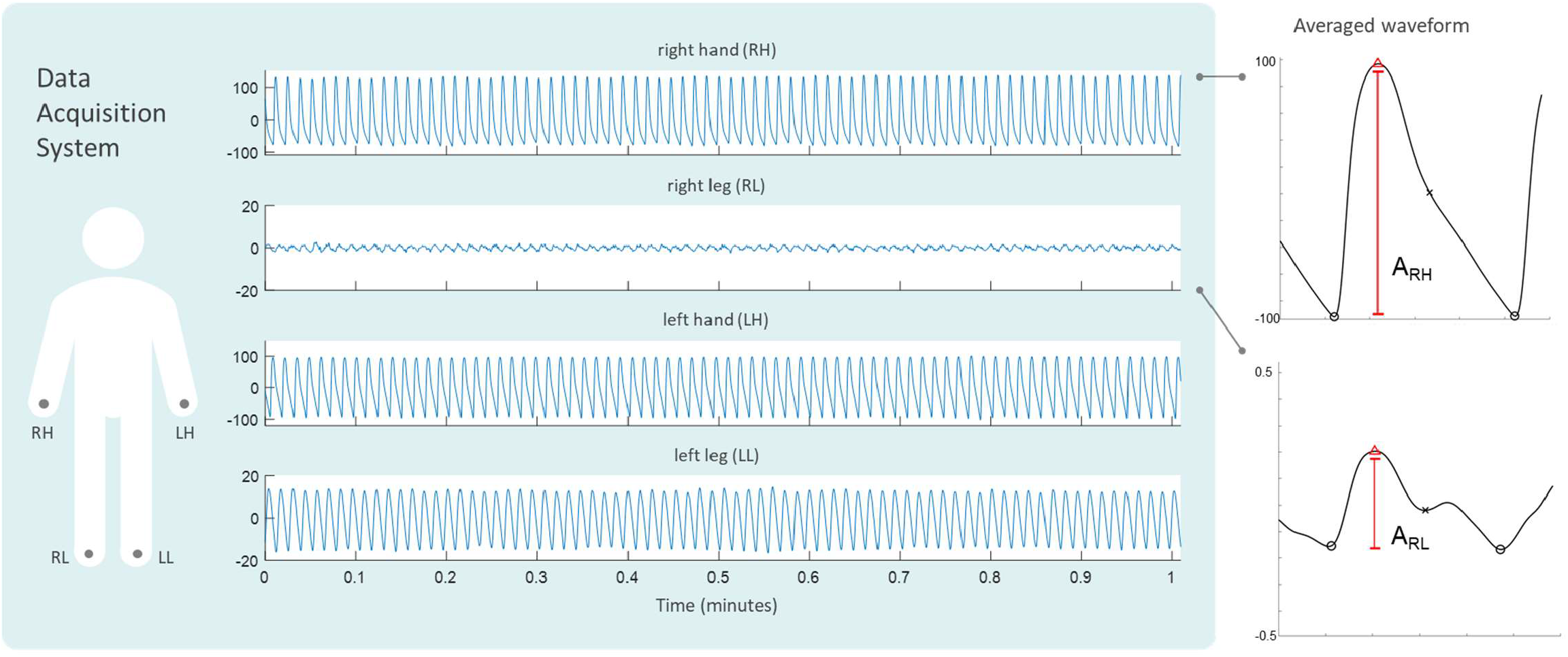
Schematic Representation of Photoplethysmography (PPG) System Implementation for PPG Signal Acquisition and Analysis. Four PPG sensors were affixed to the distal extremities of the study participants, specifically positioned on both middle fingers and second toes. Continuous PPG waveforms were simultaneously recorded from all four extremities for a minimum duration of 5 minutes. Subsequently, two key parameters were extracted from the averaged beat waveforms by aligning the PPG peaks in each location. PPG amplitude (PPG_amp_)is the amplitude of each lower extremity (e.g. A_RL_) and PPG amplitude ratio (PPG_ratio_) is calculated by the ratio of the amplitude of each lower extremity to the greater amplitude of both upper extremities (e.g. A_RL_/A_RH_).

#### PPG Amplitude (PPG_amp_)

This parameter was determined as the averaged 5-minute beat-by-beat pulse amplitude of each lower extremity.

#### PPG Amplitude Ratio (PPG_ratio_)

This parameter was calculated as the ratio of the averaged 5-minute beat-by-beat pulse amplitude of each lower extremity to the greater averaged 5-minute beat-by-beat pulse amplitude of both upper extremities.

### PAOD Diagnosis via Duplex Sonography

For vascular assessment, B-mode morphology and continuous wave Doppler measurements were conducted bilaterally on the following arteries-bilateral radial artery, ulnar artery, external iliac artery (EIA), common femoral artery (CFA), superficial femoral artery (SFA), deep femoral artery, popliteal artery (Pop A), anterior tibial artery (ATA), DPA, peroneal artery, PTA, and medial and lateral plantar artery. Duplex sonography findings were interpreted independently by two cardiologists, both of whom adhered to the University of Washington Duplex criteria. Agreement was reached between the two cardiologists. Only lesions classified as 50-99% stenotic or total occlusion were considered indicative of PAOD.

### Statistical analysis

Descriptive statistics were presented as mean ± standard deviation for continuous variables and as proportions for categorical variables. The performance of diagnostic tests for PAOD was assessed in terms of accuracy, sensitivity, specificity, and receiver operating characteristic (ROC) analysis. [16] Optimal diagnostic cut-points were determined using the Youden Index from ROC curves. Multivariate logistic regression models were developed for both ABI and PPG-derived parameters. To evaluate the discriminative ability of these models, the area under the ROC curve (AUC) was computed. ROC curves for various parameters and multivariate models were compared using the DeLong method. Statistical analyses were carried out using IBM SPSS Statistics for Windows (version 26; IBM Corp., Armonk, NY, USA) and R software (Version 3.5.0).

## Results

### Participant Demographics

From October 24, 2019, to October 12, 2021, a total of 130 participants aged between 40 and 85 years (92 men and 38 women), constituting 259 lower limbs (with one prosthetic implant case excluded), were enrolled in the study. The demographics of the current study population are summarized in Table 1. The mean age was 68.1±9.9 years (ranging from 40 to 85 years). The prevalence of risk factors and comorbidities included diabetes mellitus (DM) (76%), hypertension (75%), dyslipidemia (80%), smoking (25%), coronary artery disease (CAD) (32%), and cerebral vascular accident (CVA) (5%). Furthermore, 36% of the study participants had a diabetes duration exceeding 10 years, while 15% were in the advanced stage (3b-4) of chronic kidney disease (CKD).

**Table 1.** Basic demographics of all study subjects.

| **Table 1.** Basic demographics of all study subjects |  |  |
| --- | --- | --- |
| Variable | Value | PAOD |
| Patient number/Lower extremity number | 130/259 <sup>a</sup> | 65(50%)/111(42.8%) |
| Male/Female | 92/38 | 48(52.2%)/17(44.7%) |
| Age (year) | 68.1±9.9 |  |
| Height (cm) | 162.6±8.2 |  |
| Weight (kg) | 66.8±10.2 |  |
| Body mass index | 25.1±3.9 |  |
| Risk factor |  |  |
| Diabetes mellitus <sup>b</sup> | 100 | 49(49%) |
| < 10 years | 48(37%) | 14(29.2%) |
| > 10 years | 52(40%) | 35(67%) |
| Hypertension <sup>b</sup> | 98(75%) | 51(52%) |
| Dyslipidemia <sup>b</sup> | 104(80%) | 52(50%) |
| Smoking | 33(25%) | 25(75.8%) |
| Comorbidity |  |  |
| Coronary artery disease <sup>c</sup> | 42(32%) | 27(64.3%) |
| Cerebral vascular accident <sup>c</sup> | 7(5%) | 6(85.7%) |
| Chronic kidney disease, stage 1-3a <sup>b</sup> | 111(85%) | 49(44.1%) |
| Chronic kidney disease, stage 3b-4 <sup>b</sup> | 19(15%) | 16(84.2%) |
| Fontaine classification of the 65 subjects with PAOD |  |  |
| Fontaine classification 1 | 45(69%) |  |
| Fontaine classification 2 | 5(8%) |  |
| Fontaine classification 3 | 2(3%) |  |
| Fontaine classification 4 | 13(20%) |  |

<sup>a</sup> One prosthetic implant
<sup>b</sup> Diabetes mellitus, hypertension, dyslipidemia, and chronic kidney disease documented from electric medical chart record with treatment
<sup>c</sup> Coronary artery disease and cerebral vascular accident documented from electric medical chart record with a coronary angiogram, computed tomography, or magnetic resonance image proof.

### Univariate and Multivariate Analysis of PAOD-Associated Variables

To identify predictors of PAOD, we initially examined several variables, including basic data [age, body mass index (BMI), and gender], risk factors (diabetes mellitus of any duration, diabetes mellitus duration exceeding 10 years, hypertension, dyslipidemia, and smoking), and comorbidities [coronary artery disease (CAD), cerebral vascular accident (CVA), and advanced chronic kidney disease (CKD)] using univariate binary logistic regression. Variables such as body mass index (P=0.264), female gender (P=0.479), diabetes mellitus of any duration (P=0.702), hypertension (P=0.267), and dyslipidemia (P=0.304) were not found to be significant predictors and were excluded from further analysis. The remaining significant variables from the univariate analysis were included in a multivariate logistic regression model. This analysis revealed that PPG_ratio_ (OR: 10.700; 95% CI: 3.392-33.751, p < 0.001) and PPG_amp_ (OR: 8.694; 95% CI: 3.308-22.849, p < 0.001) were independently associated with PAOD after adjusting for relevant clinical variables. Additionally, the presence of ABI<0.9 (OR: 16.756; 95% CI: 2.722-103.166, p = 0.002), diabetes mellitus duration exceeding 10 years (P < 0.001, OR: 2.839, 95% CI: 1.547–2.212), and a history of cerebral vascular accident (OR: 8.283; 95% CI: 1.948–35.223, p = 0.004) remained independent factors associated with PAOD in the multivariate model (Table 2).

**Table 2.** Univariate and Multivariate Logistic Regression Models for Each Predictor Variable.

|  |  |  |  |  |  |  |
| --- | --- | --- | --- | --- | --- | --- |
| Predictor Variables | Unadjusted OR | 95% CI | P | Adjusted OR | 95.0 % CI | P |
| Age (1-year increase) | 1.049 | 1.020-1.079 | 0.011 |  |  |  |
| BMI | 1.038 | 0.973-1.107 | 0.264 |  |  |  |
| Gender: female | 0.821 | 0.476-1.146 | 0.479 |  |  |  |
| DM with any duration | 0.893 | 0.499-1.596 | 0.702 |  |  |  |
| DM duration >10 years | 2.839 | 1.547-2.212 | **0.001** | 3.038 | 1.155-7.987 | **0.024** |
| HTN | 1.395 | 0.775-2.509 | 0.267 |  |  |  |
| Dyslipidemia | 1.389 | 0.742-2.602 | 0.304 |  |  |  |
| Smoking | 3.761 | 2.082-6.795 | **<0.001** |  |  |  |
| Advanced CKD | 6.189 | 2.702-14.177 | **<0.001** |  |  |  |
| CAD | 2.856 | 1.668-4.893 | **<0.001** |  |  |  |
| CVA | 5.317 | 1.446-19.543 | **0.012** | 15.795 | 2.081-119.863 | **0.008** |
| ABI<0.9 | 92.367 | 21.780-391.723 | **<0.001** | 16.756 | 2.722-103.166 | **0.002** |
| PPG<sub>ratio</sub> | 38.747 | 18.269-82.180 | **<0.001** | 10.700 | 3.392-33.751 | **<0.001** |
| PPG<sub>amp</sub> | 24.612 | 12.715-47.640 | **<0.001** | 8.694 | 3.308-22.849 | **<0.001** |

### Sensitivity, Specificity, and Accuracy of PPG-Derived Parameters vs. ABI

Out of the 130 subjects in the study, 65 were diagnosed with PAOD. Among the 259 lower extremities examined, 111 were diagnosed with PAOD by duplex sonography. Specifically, 11 cases (10%) involved above-knee arteries (4 EIA and 7 SFA and/or Pop A), 79 cases (71%) involved below-knee arteries (37 ATA, 10 PTA, and 32 both ATA and PTA), and 21 cases (19%) involved both above-knee and below-knee arteries. The AUCs of ABI, PPG_ratio_ and PPG_amp_ are 0.82, 0.88, and 0.90 respectively, indicating good discriminatory performance. The ROC analysis revealed optimal cut-off values of 0.417 for PPG_ratio_ and 58 for PPG_amp_ in diagnosing peripheral arterial obstructive disease (PAOD).

Both PPG_ratio_ and PPG_amp_ with the optimal thresholds (PPG_ratio_ <0.417 and PPG_amp_ <58) demonstrated higher sensitivity levels, 78.4% (95% CI: 70.3%-85.6%; p < 0.05) and 75.7% (95% CI: 67.6%-82.9%; p < 0.05), in comparison to ABI<0.9, which had a sensitivity of 55.9% (95% CI: 46.6%-65.8%), while the specificity of PPG_amp_ [92.6%; 95% CI:87.8%-96.6%] was not differ to ABI<0.9 [98.7%; 95% CI:96.6%-100%]. The overall accuracy of ABI<0.9, PPG_amp_, and PPG_ratio_ were 80.3% (95% CI: 76.5%-84.6%), 85.3% (95% CI: 81.1%-89.2%), and 83.4% (95% CI: 78.8%-87.6%), respectively (Figure 2A).

**Figure 2.**
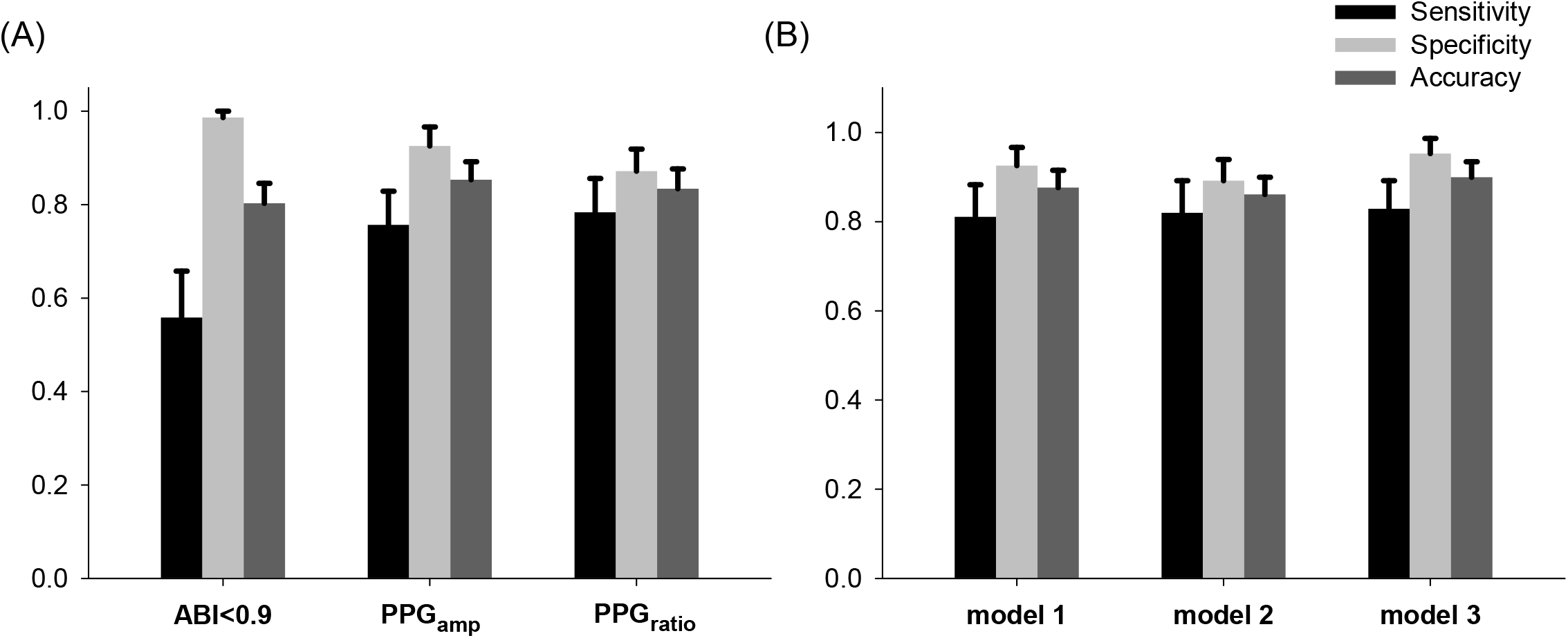
(A) The sensitivity, specificity, and accuracy of the ABI<0.9 and PPG derived parameters with optimal threshold for diagnosis of PAOD and their 95% confidence intervals. (B) The combination models with ABI<0.9 and PPG derived parameters with optimal threshold and their 95% confidence intervals.

Among the 111 cases diagnosed with PAOD by duplex sonography, 62 (55.9%) have an ABI < 0.9, 87 (78.4%) have a PPG_ratio_ < 0.417, 84 (75.7%) have a PPG_amp_ < 58; 42 (37.8%) of the 111 cases have an 0.9< ABI< 1.3, yet 28 in the 42 (25.2%) have a PPG_ratio_ < 0.417 and 25 in the 42 (22.5%) have a PPG_amp_ < 58; 7 (6.3%) of the 111 cases have an ABI> 1.3, yet 4 in the 7(3.6%) have a PPG_ratio_ < 0.417 and 4 (3.6%) in the 7 have a PPG_amp_ < 58.

### Using Combined Models of ABI and PPG-derived Parameters

The combination models, including model 1 (ABI<0.9 and PPG_ratio_), model 2 (ABI<0.9 and PPG_amp_), and model 3 (ABI<0.9, PPG_ratio_, and PPG_amp_), exhibited improved performance compared to using ABI alone, with AUCs of 0.922, 0.922, and 0.931, respectively (Delong test p < 0.01) (Figure 3). The sensitivity, specificity, and accuracy after the combination were 81.1% (95%CI: 73.9%-88.3%), 92.6% (95%CI: 88.6%-96.7%), and 87.6% (95%CI: 83.8%-91.5%) for model 1, 82.0% (95%CI: 74.8%-89.2%), 89.2% (95%CI: 84.5%-93.9%), and 86.1% (95%CI: 81.9%-90.0%) for model 2 and 82.9% (95%CI: 75.7%-89.2%), 95.3% (95%CI: 91.9%-98.7%), and 90.0% (95%CI: 86.1%-93.4%) for model 3 (Figure 2B). The sensitivity and accuracy of model 3 were significantly higher compared to ABI<0.9 alone (p<0.05).

**Figure 3.**
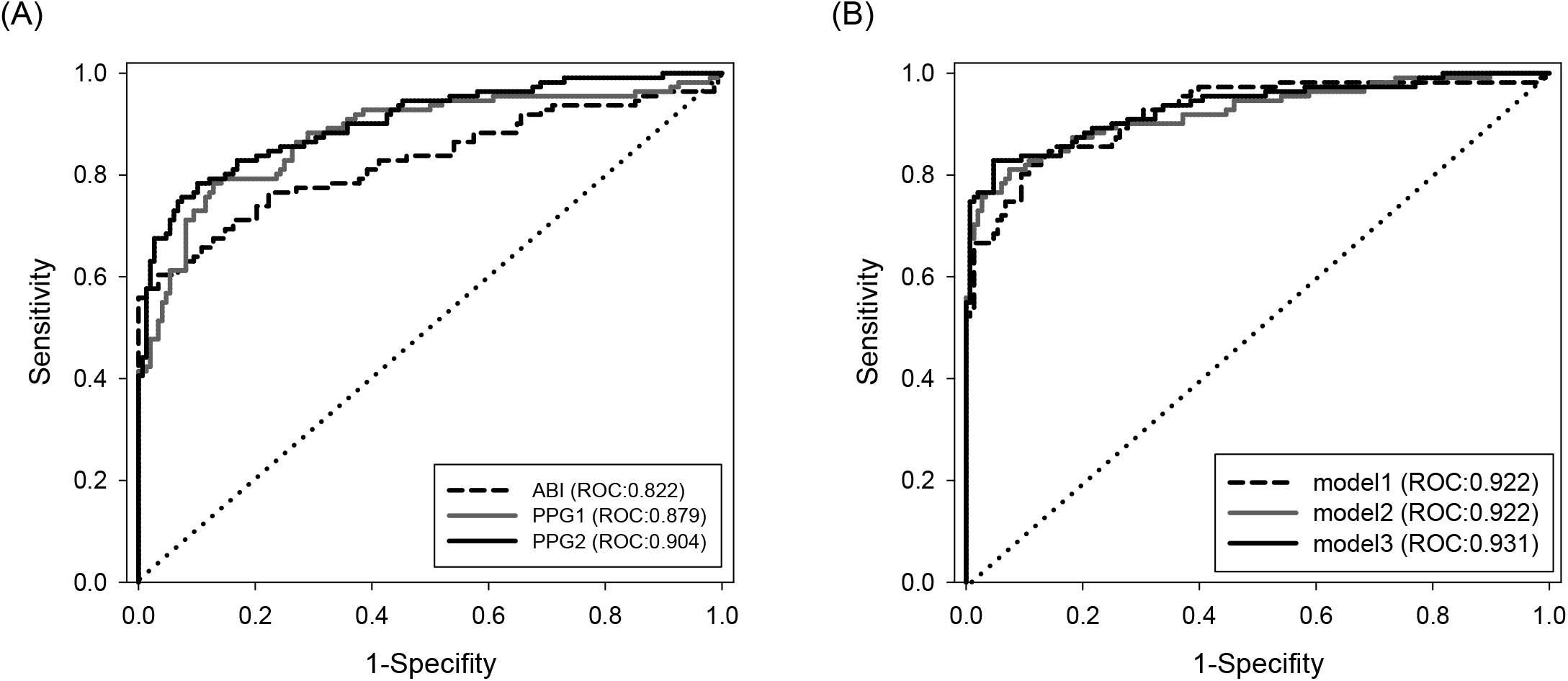
Receiver operating characteristic (ROC) curves comparing the diagnostic performance of the ABI, PPG_ratio_, and PPG_amp_ for PAOD diagnosis. (A) ROC curves for each diagnostic test. (B) ROC curves for logistic regression models using different combinations of parameters: Model 1 (ABI>0.9 and PPG_ratio_); Model 2 (ABI>0.9 and PPG_amp_); and Model 3 (ABI>0.9, PPG_ratio_, and PPG_amp_). The area under the curve (AUC) is presented for each curve as a measure of diagnostic accuracy.

## Discussion

This study systematically examined the sensitivity, specificity, and accuracy of ABI, parameters derived from PPG, and combinations in diagnosis of PAOD based on duplex sonography. First, PPG demonstrated higher sensitivity compared to ABI, particularly in individuals with a high atherosclerotic risk profile. Second, ABI’s sensitivity was notably lower at the below-knee level, especially in cases of single stenotic or occluded ATA or PTA. This discrepancy may be attributed to both medical sclerosis and the design of the ABI measurement. Third, combining ABI and PPG parameters can serve as a simple yet effective method for enhancing sensitivity and accuracy in the diagnosis of PAOD.

### The Influence of PAOD Locations on ABI Sensitivity

The ABI serves as the primary worldwide noninvasive diagnostic tool recommended for PAOD screening. However, its sensitivity is inadequate in specific cases, such as individuals with diabetes, advanced CKD, advanced age, or those on long-term immunosuppressive therapy, primarily due to the occurrence of medial sclerosis. In such situations, measuring the great toe pressure and calculating the TBI is recommended, especially when ABI exceeds 1.3 or when patients have incompressible ankle arteries.[11, 12] Similar to ABI, TBI is determined by comparing the systolic blood pressure in the great toe to that in the arm. This involves using arm cuffs and toe cuffs for pressure measurement but the diagnostic cutoff value for a pathologic TBI is less than 0.7. However, TBI primarily assesses perfusion in the PTA and medial plantar artery. It does not directly evaluate the circulation in the lateral part of the forefoot, which is supplied by the DPA and ATA. Furthermore, the threshold of 0.7 is also not well-proven scientifically.[17] The treadmill exercise test combined with ABI is also recommended for symptomatic patients with normal ABI. However, arranging and performing this test is time-consuming, and certain contraindications restrict its use.[18, 19]

The ABI showed variations depending on the stenotic or occluded levels. In our study, the sensitivity of ABI was lower in the below-knee level compared to the above-knee level. Furthermore, when examining below-knee levels with single artery stenosis/occlusion, ABI demonstrated notably low sensitivity (19.1%). Among the 111 stenotic/occluded arteries, 47 were single stenotic/occluded arteries in the below-knee level, including 37 ATA and 10 PTA. When these cases were excluded from the analysis, the overall sensitivity of ABI in the remaining 64 stenotic/occluded arteries increased from 55.9% to 83.1% based on duplex sonography results. This phenomenon can be attributed to the calculation formula of the ABI, which typically involves measuring the systolic blood pressure of both brachial arteries, DPA, and PTA above the malleolus and the ABI is calculated using the higher systolic pressure of either PTA or DPA for each lower extremity as the numerator and the higher of both brachial artery systolic pressures as the denominator. In a study by Frank Schröder et al., they proposed a modified calculation of ABI with the lower systolic pressure of PTA or DPA as the numerator. Their findings indicated that this modification elevated the sensitivity of ABI from 68% to 89%, albeit with a moderate decrease in specificity (from 99% to 93%) compared to conventional ABI. They suggested that medial calcinosis of PTA or DPA might lead to overestimation of lower limb pressure, resulting in low sensitivity in conventional ABI.[20] In our study, among the 47 single stenotic/occluded arteries, 34 were occluded with collateral flow and exhibited lower systolic pressure, while the remaining 13 were 50-99% stenotic. We believe that, in addition to medial calcinosis, the choice of the “higher systolic pressure of PTA or DPA” as the numerator in ABI measurements might prioritize the healthier vessel with higher systolic pressure and neglect the occluded vessel with significantly lower systolic pressure, contributing to the observed low sensitivity of ABI.

In our study, we employed four PPG sensors similar to the cuffs of TBI but simplified our procedure by directly measuring volume changes peak instead of pressure. This eliminates the need for cuff inflation and deflation. Additionally, our choice of PPG sensor placement at the second toe allows us to assess both distal perfusions, from PTA to the medial plantar artery and ATA to DPA, providing a more comprehensive evaluation.

### Improvement of diagnostic sensitivity with PPG in patients at high atherosclerotic risk

The prevalence of disease or associated risk factors can significantly influence the sensitivity of diagnostic tools. In a comprehensive review by Dachun Xu et al. on the sensitivity and specificity of ABI, they reported high specificity (83.3–99.0%) and accuracy (72.1–89.2%) when ABI was less than 0.9 in PAOD. However, they showed a relatively low sensitivity (15–79%) in patients of advanced age or those with diabetes.[11]

In our study, the subjects had an average age of 68.1±9.9, with 78% of them having diabetes mellitus. Notably, both PPG_ratio_ and PPG_amp_ demonstrated superior overall sensitivity and accuracy compared to ABI in this particular conditions. Additionally, PPG parameters exhibited improved sensitivity in subjects with other atherosclerotic risk factors such as hypertension (HTN), dyslipidemia, chronic kidney disease (CKD), and smoking (supplementary table 1). However, it is important to highlight that ABI still maintained very high overall specificity.

Diabetes mellitus is associated with chronic inflammation, which can induce atherosclerosis and accelerate its progression. This inflammatory response contributes to the elevated mortality and morbidity rates associated with cardiovascular disease. [21] The duration of diabetes was also identified as a prominent factor associated with macrovascular events or death, as indicated by the results of the Action in Diabetes and Vascular Disease: Preterax and Diamicron Modified Release Controlled Evaluation (ADVANCE) trial. [22] Based on the Framingham Heart Study, Caroline SF, et al, reported a 1.86 times higher risk of coronary heart disease death for each 10-year increase in diabetes duration.[23]

In our study, ABI, PPG_ratio_ and PPG_amp_ are independently associated with PAOD after adjusting for variables such as smoking, age, advanced CKD, CVA, and CAD risk factors, and mellitus duration exceeding 10 years. In addition, many of these individuals were asymptomatic. This underscores the potential utility of combining PPG and ABI for PAOD screening in this specific group.

### Potential Implications of PPG-Derived Parameters as a Complementary Tool to ABI

In a study by Allen J. et al., they reported diagnostic parameters for PPG using the shape index method, with an 88.9% sensitivity, 90.6% specificity, and 90.2% accuracy in diagnosing PAOD. Notably, their study referenced the established Ankle-Brachial Index (ABI) measurement results. [13, 14] In contrast, our PPG study was conducted with reference to duplex sonography results, enabling us to directly compare the diagnostic performance of PPG with that of ABI. The results from our study suggest that PPG can significantly enhance the sensitivity of ABI, particularly in cases involving below-knee lesions and individuals with atherosclerotic risk factors (Supplementary table 1). Similar to ABI preparation, participants in our study would recline on an examination table in a warm environment for 5-10 minutes before the test. The advantage of PPG lies in its ability to be performed nearly simultaneously with ABI assessment using PPG sensors, a simple, cost-effective, and non-invasive optical tool. In addition to traditional methods such as TBI or the treadmill exercise test, the combination of ABI and PPG offers a viable alternative for the early detection of PAOD, especially in patients at high risk of atherosclerosis. When combining either PPG_ratio_ or PPG_amp_ with ABI, our study demonstrated improved sensitivity and accuracy compared to relying solely on PPG or ABI.

## Limitations

Our study has some limitations. Firstly, our study was performed at room temperature between 23 to 25°C. A pre-heating infrared heater had been used if the temperature was less than 23°C. The study result could not fit the condition with temperatures less than 23°C or more than 25°C. Secondly, our study design excluded the subjects with CKD stage V in uremic status who were known to have Mönckeberg’s medial sclerosis.[24] Thirdly, ABI and TBI have a U shape and linear association with cardiovascular morbidity respectively. Our study group contained a relatively small sample size (130 subjects with 259 lower extremities). A further comprehensive study with a larger sample size was warranted to evaluate the ABI and PPG combination’s reproducibility and the association with cardiovascular morbidity or mortality.

## Conclusion

This study demonstrated that ABI has lower sensitivity than PPG in the detection of PAOD in participants with a high risk of atherosclerosis or below-knee single stenotic/occluded artery. The combination of ABI and PPG could improve the sensitivity and accuracy prominently in diagnosing PAOD compared to ABI or PPG alone. In patients with high atherosclerotic risks, such as DM duration exceeding 10 years or advanced age, the combination of ABI and PPG could provide an alternative simple screening method that can prompt further investigation of the need for more advanced imaging studies.

## Data Availability

Data will be available upon reasonable request, conditional upon obtaining Institutional Review Board (IRB) approval

## Author contributions

H.L.H., and C.L. designed the study. H.L.H., and C.C.H. acquired the samples, clinical outcomes, and carried out the experimental work. H.L.H., M.T.L., and C.L. carried out the analysis. H.L.H wrote the paper, which was edited by C.L. All authors read and approved the manuscript.

